# Humans with Pulmonary Arterial Hypertension display a global hypermethylation signature that worsens in patients who have a mutation in the gene encoding the methylation eraser, Tet Methylcytosine Dioxygenase 2 (TET2)

**DOI:** 10.1101/2023.08.09.23293846

**Authors:** Charles C.T Hindmarch, François Potus, Al-Qazazi Ruaa, Brooke Ring, William C Nichols, Michael J Rauh, Stephen L Archer

**Affiliations:** Department of Biomedical and Molecular Science (DBMS), Queen’s University, Kingston, Ontario; Department of Medicine, Queen’s University, Kingston, Ontario; Queen’s CardioPulmonary Unit, Translational Institute of Medicine, Department of Medicine, Queen’s University, Kingston, Ontario, Canada; Pulmonary Hypertension Research Group, Center de Recherche de l’Institut Universitaire de Cardiologie et de Pneumologie de Québec, QC, Canada; Division of Human Genetics, Cincinnati Children’s Hospital Medical Center, and Department of Pediatrics; University of Cincinnati College of Medicine, Cincinnati, Ohio, USA; Department of Pathology and Molecular Medicine, Queen’s University, Kingston, Ontario, Canada

**Keywords:** Pulmonary arterial hypertension (PAH), Clonal hematopoiesis of indeterminate potential (CHIP), *Metabolism*, *Inflammation*, Immune, cell differentiation, DNA methylation

## Abstract

Epigenetic changes in gene expression due to DNA methylation are important physiologic and pathologic regulators of pulmonary vascular structure and function. Genetic or acquired alterations in DNA methylation or demethylation have been associated with the development and progression of pulmonary arterial hypertension (PAH). However, the DNA methylome signature of human PAH and its consequences on the PAH transcriptome are unknown. Reduced Representation Bisulfite Sequencing (RRBS) was used for epigenome-wide mapping of DNA methylation in whole peripheral blood of 10 healthy people and 20 age/sex matched PAH patients. RNA deep sequencing was performed in parallel on the same samples. We used whole-exome sequencing to identify two PAH cohorts, one free of mutations in the genes know to be associated with PAH patients and the second had mutations of *TET2*, a newly identified PAH gene that encodes an enzyme that mediates DNA demethylation. We report an increased in global DNA methylation in the blood of PAH patients compared to healthy controls. Patients carrying the *TET2* mutation had a further increase in DNA compared to mutation free PAH patients. We identified 1,069 unique Differentially Methylated Regions (DMR) in the blood of PAH patients with a TET2 mutation. When organized into functional groups, we observed an enrichment of genes involved in ‘immune’, ‘cell differentiation’, or ‘metabolic’. When we compared these genes to publicly available data from an independent study on blood from PAH patients compared to controls, we identified 218 mRNA transcripts that align with our data (e.g. are hypermethylated blood genes that are downregulated in PAH blood), and functional analysis of these genes reveals common enriched terms in ‘immune’, ‘cell differentiation’, or ‘metabolic’ function. We characterized DNA methylation changes associated with TET2 mutation in the blood of patients with PAH compared to controls. This data demonstrates that epigenetic regulation of genes by methylation is involved in altered immune function, cell differentiation and metabolism in PAH.

## Introduction

Pulmonary arterial hypertension (PAH) is a lethal vasculopathy characterized by obliterative remodelling, stiffening and vasoconstriction in the pulmonary arterial circulation. Although 18 PAH genes have been identified, mutations in known genes only occur in ∼20% of idiopathic cases versus ∼70% in familial cases. Most PAH mutations have an autosomal dominant mechanism of inheritance with variable penetrance. Epigenetic regulation represents a mechanism for molecular tuning of genome expression, by which environmental factors modulate gene transcription but not genetic code. DNA methylation, facilitated by the DNA methyltransferases (DNMT), which adds methyl group to CpG sites, inhibits gene transcription by interfering with the access of transcription factors to promoter and enhancer regions. Conversely, TET2 catalyzes the removal of methyl groups. Recently, we^1^ and others^2^ demonstrated that *TET2* mutations are a novel cause of PAH.

To date, no comprehensive analysis of whole human DNA methylome or an assessment of the potential compounding effects of *TET2* mutation on DNA methylation in PAH has been performed. We predicted that loss of function mutations of TET2 would cause a panchromosomal increase in DNA methylation. Here, we identify a global hypermethylation in the blood of patients with PAH, that is exacerbated by inactivating *TET*2 mutations.

Patient samples were obtained following informed consent, approved by Institutional Review Boards (#6016826). Clinical characteristics are described in **Figure1a**. We assessed genome-wide DNA methylation in blood obtained from 20 PAH patients and 10 age/sex-matched healthy participants (mean age 72 years, 70% female). The PAH cohort includes 10 PAH patients carrying deleterious *TET2* mutations^1^ and 10 without mutation in *TET2* or other established PAH genes (*BMPR2; EIF2AK4; TBX4; ATP13A3; GDF2; SOX17; SMAD4; SMAD1; KLF2; BMPR1B; KCNA5; AQP1; ACVRL1; SMAD9; ENG; KCNK3; CAV1*)^3^. PAH cohorts were matched for disease severity and include 60% IPAH and 40% APAH. While we observed a 0.776% increase in methylation in PAH patients, there was a 1.124% increase in methylation in the *TET2* group compared to healthy controls. At the chromosome level, significant hypermethylation in the PAH patients without mutation is restricted to specific chromosomes (Chr 1, 10, 12, 16, 2, 22, 5, 7, 8, 9 and Y). However, in patients with PAH and a *TET2* mutation, significant changes in methylation are observed on every chromosome (**Figure1b**). We clustered single CpG sites to define larger differentially methylated regions (DMR) in the patients with *TET2* mutation and observed 2,520 DMR, of which 2,333 are hypermethylated, and 187 hypomethylated. Statistical analysis (areaStat score >10) revealed 1,069 unique DMR within annotated protein coding genes (245 Promoter, 575 Exon, 1,325 Intronic; some targets overlap; **Figure1c**). Functional gene ontology (GO) analysis of these genes reveals 312 significant terms including a cluster of terms related to either ‘immune’, ‘differentiation’ or ‘metabolic’ revealing a functional profile of genes involved in these biological processes (**Figures1d/e/f**). We compared our DMRs to work linking TET2 associated CpGs to coronary-artery disease^4^, and observed general overlap with our function-organized; specifically for leukocyte activation (GO:0045321) and lymphocyte activation (GO:0046649). We wanted to identify if these DMRs impact gene expression, so we remined public data (GSE38267)^5^ from PAH patient blood compared to healthy blood. In total, we identify the common downregulation of 218 transcripts in that independent dataset which align with our putative DMRs (i.e., hypermethylated DMRs and associated transcript downregulation). When GO-analysis was performed on these transcript-filtered DMRs, many similar targets involved in ‘metabolism’, ‘immune’ and cell ‘differentiation’ were observed (**see numbers in parenthesis in Figure 1D/E**).

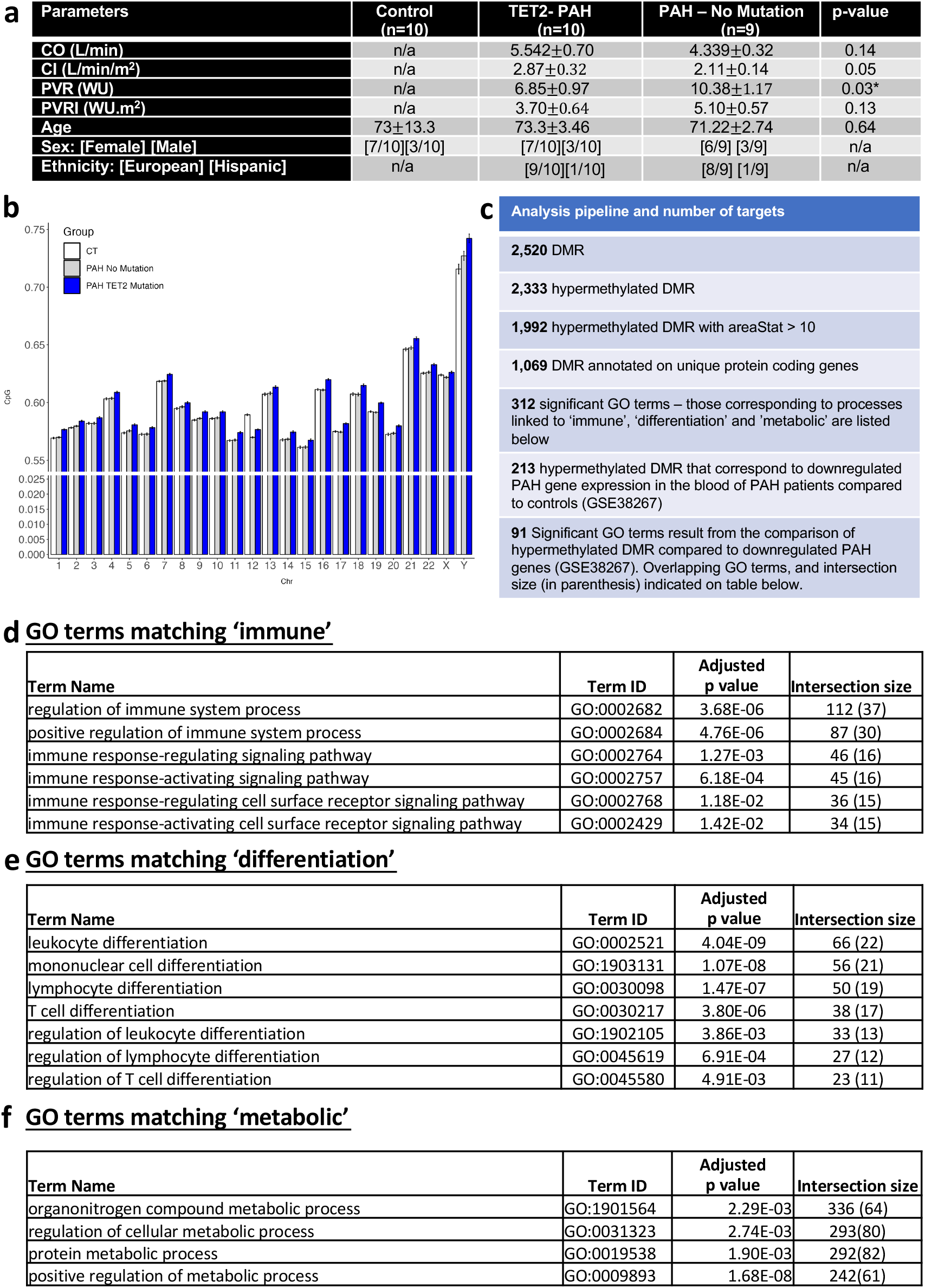

Here, we have demonstrated that the blood from PAH patients is characterized by a global hypermethylation, which is exacerbated by the presence of a deleterious *TET2* mutation (because a major mechanism to remove methyl groups is impaired). This study was performed on whole blood, and we do not know whether these DMR also manifest in the lung, pulmonary artery, and the right ventricle of the heart; however, in our prior work, deletion of *TET2* in hematopoietic cells was sufficient to cause PAH in mice^1^. Intriguingly, the hypermethylated DMRs occur in pathways that are already highly implicated in PAH pathogenesis; cell differentiation, immunity, mitochondrial metabolism and inflammation. We hypothesize: **1**. that there is a causal link between *TET2* mutation and panchromosomal gene hypermethylation; **2**. there may decreased expression of critical genes in PAH-relevant GO pathways involving metabolism, inflammation/immunity, and cell differentiation; **3**. the epigenetic signal relevant to *TET2* mutation is identifiable in peripheral blood cells, suggesting it as a putative and clinically accessible biomarker. More work is required to identify which genes are dysregulated as a direct consequence of a mutation in *TET2* and to determine if these mutations occur primarily in the hematopoietic system, as in clonal hematopoiesis of indeterminate potential (CHIP) or in the cells of the cardiopulmonary system. The occurrence of hypermethylated DMRs even in PAH patients lacking *TET2* mutations suggests unidentified epigenetic mechanism exist.

## Data Availability

Aggregate data can be shared with researchers at established Universities or Medical schools upon request.

## Reviewer comments

### Reviewer #1

In this letter the authors show that in 20 patients with PAH there is an increase in the methylation of DNA in most chromosomes. Of those patients, in 10 that had loss of function mutations there was further DNA methylation that included genes regulating metabolism and inflammation (both relevant to PAH). Along with their previous work in mice showing a potentially causal effect of TET2 loss in rodent PHT models, this letter provides further insight on genetic causes of human PAH.The letter is well written and clear. The information provided is novel and would be of interest to the readers of Circulation. The epigenetic basis of PAH is rapidly being revealed and holds promise for the generation of novel biomarkers and therapies. The authors need to clarify how the 10 patients with and the 10 without TET2 mutations were selected (where they consecutive referrals or selected? It would be helpful to have some idea of the incidence of these mutations in the PAH population). The authors also need to clarify that in their cohort, there was no evidence of CHIP/myelodysplasia. Since the incidence of CHIP increases over the age of 60 and their TET2 cohort was 73 years old, it would be important to know whether there is a possibility of CHIP and PAH coexisting. In the absence of CBC values, a simple statement that there was no clinical evidence of myelodysplasia would suffice.

### Reviewer #2

In this manuscript, the authors performed DNA methylome analysis of blood cells from PAH patients (with or without TET2 mutation) and control subjects. They found that there is an increase in methylation in PAH patients, compared to healthy controls. Methylation was further increased in PAH patients with TET2 mutation, compared to blood cells from PAH patients without TET2 mutation. Analysis of DMR in PAH with TET2 mutation indicated that pathways associated with PH are altered in their DNA methylation. The role of TET2 mutation in increasing expression of inflammatory genes in hematopoietic cells are well-established. In addition, the involvement of hematopoietic cells in PH development and progression are also have been extensively reported. Unfortunately, the study has been focusing on DNA methylation alteration in blood/hematopoietic cells, but not pulmonary vascular wall cells. The analysis of the role of TET2 mutation in pulmonary vascular wall cells becomes more urgent because the authors (Circulation, 2020) have reported that PH patients with TET2 mutation (even in germline) exhibit older mean age of onset (66.9 years versus 18 years in APAH+IPAH) and lower mPAP (43.3 mmHg versus 50.0 mmHG in APAH+IPAH), which indicate that the function of TET2 mutation could be opposite in hematopoietic cells (TET2 mutation leads to increased inflammation) and in pulmonary vascular wall cells (TET2 mutation leads to suppression of inflammation and/or other processes). Other minor concerns are below:

1. Recently, we and others demonstrated that TET2 mutations are a novel cause of PAH. This is an overstatement as TET2 mutation in hematopoitic cells, but not in germline or whole body, leads to PH onset.
2. Result of DMR on PH patients without TET2 mutation versus controls or versus PH patents with TET2 mutation, should also be analyzed and presented.
3. DNA methylation analysis data need to be deposited if accepted.

